# When Conventional Methods Fail: First Detection of a *Candida viswanathii* Outbreak in Europe in a Paediatric Hospital Revealed by Whole Genome Sequencing and FT-IR Spectroscopy

**DOI:** 10.1101/2025.10.25.25337813

**Authors:** Gianluca Vrenna, Valeria Fox, Venere Cortazzo, Serena Raimondi, Marco Cristiano, Gianluca Foglietta, Sara Carilli, Martina Rossitto, Barbara Lucignano, Manuela Onori, Maria Paola Ronchetti, Andrea Dotta, Andrea Campana, Lorenzo Galletti, Luca Di Chiara, Alberto Villani, Marta Luisa Ciofi Degli Atti, Daniela Perrotta, Corrado Cecchetti, Massimiliano Raponi, Carlo Federico Perno, Paola Bernaschi

**Author notes:** **Correspondence:** Venere Cortazzo. These authors contributed equally to this work and share first authorship (Gianluca Vrenna, Valeria Fox). These authors contributed equally to this work and share last authorship (Carlo Federico Perno, Paola Bernaschi).

## Abstract

**Objectives:** *Candida viswanathii* has been sporadically reported in Asia and South America but never in Europe. This study reports the first European outbreak of *C. viswanathii* in a paediatric hospital, outlining diagnostic challenges and containment measures.

**Methods:** Fifteen *C. viswanathii* isolates were recovered from blood cultures of consecutive paediatric patients admitted to intensive care units between April and August 2025. Identification was performed using MALDI-TOF MS, chromogenic media, Fourier-transform infrared (FT-IR) spectroscopy, and whole-genome sequencing (WGS). Antifungal susceptibility testing was performed by broth microdilution.

**Results:** All isolates were initially misidentified as *C. tropicalis* by MALDI-TOF MS and undetected by the FilmArray BCID2 panel. WGS confirmed *C. viswanathii*, and FT-IR analysis revealed clonally related strains, indicating an outbreak. Colonies displayed a distinct deep-blue colour on chromogenic medium. Elevated fluconazole minimum inhibitory concentrations were observed, while isolates remained susceptible to echinocandins and amphotericin B. A multidisciplinary infection-control response halted transmission within four weeks.

**Conclusions:** This investigation documents the first *C. viswanathii* outbreak in Europe, highlighting diagnostic limitations of current commercial tools and the need for updated databases. Integration of FT-IR spectroscopy and WGS facilitated outbreak detection and containment, underscoring the importance of advanced diagnostics and surveillance for emerging fungal pathogens.

## Introduction

Human-pathogenic Candida species exhibit considerable biological heterogeneity and variable susceptibility to antifungal agents, features particularly relevant in pediatrics where defining optimal therapeutic strategies is challenging. Accurate species-level identification, including of non-albicans Candida (NAC), remains a critical issue in clinical mycology [1]. This difficulty is compounded by the emergence of uncommon species, often absent from reference databases routinely employed in microbiology laboratories.

Invasive fungal infections, especially those caused by Candida, are a major source of morbidity and mortality worldwide. About 90% of invasive candidiasis cases are attributed to five species *C. albicans, C. glabrata, C. tropicalis, C. parapsilosis*, and *C. krusei*. Nonetheless, over 15 less common species have been implicated in human disease [2]. These rare pathogens differ substantially in virulence, antifungal susceptibility, and epidemiology. Consequently, rapid and accurate identification at the species level is essential, enabling the shift from empirical and potentially suboptimal therapy to targeted antifungal regimens, thereby improving outcomes [2].

Among the uncommon yeasts, *Candida viswanathii* warrants particular attention. First described in India in 1959 [3], it has been rarely reported in hospital settings. The largest case series, published in 2018 from India, described 23 bloodstream infections over four years. Additional sporadic cases were reported in Singapore, China, and Argentina, but overall descriptions remain scarce and no cases had been documented in Europe [1]. Despite its rarity, *C. viswanathii* is increasingly recognized as an opportunistic pathogen capable of causing severe and sometimes fatal infections.

A key feature of *C. viswanathii* is its frequent elevated minimum inhibitory concentrations (MICs) to fluconazole, which increase the risk of treatment failure when inappropriate antifungal therapy is administered. Such resistance may lead to persistent colonization, invasive disease, and facilitate nosocomial spread, potentially triggering outbreaks difficult to control [1]. These features underscore the need to combine precise species identification with antifungal susceptibility testing, especially in high-risk settings such as neonatal and pediatric intensive care units (NICU and PICU). While final therapeutic decisions remain clinical, early communication of resistance patterns at the time of diagnosis can guide management in line with pediatric recommendations [4,2].

Our recent experience illustrates the diagnostic challenges. *C. viswanathii* is often misidentified as *C. tropicalis* by matrix-assisted laser desorption ionization–time of flight mass spectrometry (MALDI-TOF MS), widely regarded as the diagnostic gold standard [5]. Discrepancies became evident when a positive blood culture yielded no identification with the FilmArray BCID2 panel, yet MALDI-TOF MS subsequently identified C. tropicalis. Whole-genome sequencing (WGS) of the index isolate confirmed *C. viswanathii*, and further discordant cases were rapidly recognized as clonal *C. viswanathii* strains using Fourier-transform infrared (FT-IR) spectroscopy.

In this study, we analyzed 15 *C. viswanathii* isolates recovered from pediatric patients at the NICU and CICU between April and August 2025. Our aims were to describe their clinical, microbiological, and antifungal susceptibility profiles, and to explore strategies to improve diagnostic accuracy. Specifically, we sought to: (i) identify MALDI-TOF MS spectral features that may distinguish *C. viswanathii* from *C. tropicalis*; (ii) evaluate chromogenic media (CHROMagar™ Candida) as a tool for rapid differentiation; and (iii) assess FT-IR spectroscopy (Bruker IR Biotyper) for its ability to recognize and cluster *C. viswanathii* once a reference spectrum is established.

## Materials and Methods

Fifteen *Candida viswanathii* isolates were recovered from blood cultures of pediatric patients admitted to the NICU (n=5), CICU (n=8), Transplant/Mechanical Assistance Unit (n=1), and Cardiology Unit (n=1) at Bambino Gesù Children’s Hospital (Rome, Italy) between April–August 2025. Blood samples were incubated in aerobic/anaerobic bottles (bioMérieux) using an automated monitoring system. Positive cultures, showing yeasts on Gram stain, were first analyzed with the BioFire® BCID2 panel; unidentified samples underwent direct MALDI-TOF MS testing. All positive bottles were subcultured on solid media at 37 °C, and colonies were re-examined by MALDI-TOF MS for species confirmation.

### FilmArray BCID2 panel testing

For all fifteen positive blood culture bottles, a 200 µL aliquot was processed using the BCID2 panel according to the manufacturer’s instructions. Testing was performed on the FilmArray Torch platform, and results were automatically generated by the FilmArray software. When no identification was provided by the BCID2 panel, conventional identification methods (MALDI-TOF MS and subculture) were applied as described above.

### Microbial identification by MALDI-TOF MS

Positive blood culture samples were processed using the Bruker Sepsityper® kit (Bruker Daltonics, Bremen, Germany), and the resulting pellets were analyzed with an MBT Smart MALDI-TOF mass spectrometer (Bruker Daltonics) operated in positive mode at a laser frequency of 200 Hz. Spectra acquisition and analysis were performed using MBT Compass HT IVD software version 5.2.330, with instrument calibration using Bruker Bacterial Test Standard (BTS) and spectral comparison against the MBT Compass HT IVD Library 2023 (version 1877017). When required, additional MALDI-TOF MS analyses were carried out on colonies obtained after short-term incubation on selective solid media. For comparison, isolates were also analyzed on two additional MALDI-TOF MS platforms available in the laboratory (Autobio Autof MS1000 and Zybio XS2600, both operated in positive mode at 60 Hz) using their respective acquisition and analysis software.

### Creation of a local MALDI□TOF MS reference spectrum for *Candida viswanathii*

Following confirmation of *C. viswanathii* by WGS for the index isolate and clustering of all subsequent isolates by FTIR spectroscopy, an internal protocol was implemented to expand the MALDI□TOF MS database for this species. Four WGS-confirmed *C. viswanathii* isolates were processed according to the Bruker “Creation of Library Entries” protocol (revision March 2020). Isolates were grown on solid media, and biomass was prepared following the manufacturer’s recommendations. Mass spectra were acquired on a Bruker LT system (60 Hz, positive mode) using AutoXecute MB_AutoX. Spectra were calibrated with Bruker Bacterial Test Standard (BTS), evaluated for quality, and processed with flexControl 1.4 and Compass flexAnalysis 1.4 software to generate Main Spectrum Projections (MSPs). The resulting MSPs were incorporated into the local Bruker Taxonomy library for subsequent use in routine identification.

### Retrospective analysis of 2024–2025 *Candida tropicalis* isolates

All blood culture isolates initially reported as *C. tropicalis* from January 2024 to July 2025 were re-evaluated using the updated local MALDI-TOF MS library and FT-IR spectroscopy. All isolates were sequenced by WGS for definitive species confirmation. This retrospective analysis aimed to assess whether earlier cases of *C. viswanathii* might have been misidentified as *C. tropicalis* before the 2025 cluster.

### Antifungal susceptibility testing

Antifungal susceptibility testing of all *C. viswanathii* isolates was performed using the Sensititre™ YeastOne ITAMYUCC antifungal susceptibility panel (Thermo Fisher Scientific, Waltham, MA, USA). Minimum inhibitory concentrations (MICs) for fluconazole, voriconazole, posaconazole, isavuconazole, echinocandins (anidulafungin, caspofungin, and micafungin), and amphotericin Bwere determined according to the Clinical and Laboratory Standards Institute (CLSI) broth microdilution method. MIC endpoints were read visually after 24 h of incubation at 35 ± 2 °C. Interpretation was performed according to CLSI clinical breakpoints or epidemiological cutoff values, when available (Table S1).

### FT-IR analysis

*C. viswanathii* isolates were subcultured on Sabouraud dextrose agar plates (Kima, Padua, Italy) and incubated for 24 h at 30 °C. Colonies from three biological replicates were suspended in 100 µL of deionized water; the standard preparation including 70% ethanol and a simplified protocol with water alone were both tested. Suspensions were vortexed twice with metal cylinders, and five technical replicates were spotted on a 96-well silicon plate and air-dried.

Spectra were acquired using an FT-IR Biotyper system (Bruker Daltonics, Bremen, Germany) with OPUS 8.2.28 software and analyzed in the 800–1300 cm□^1^ range using IR Biotyper software (v3.1.2). For each isolate, 15 spectra (3 biological × 5 technical replicates) were processed. Data analysis included Principal Component Analysis (PCA), Linear Discriminant Analysis (LDA), and hierarchical clustering (Euclidean distance, average linkage) to explore relatedness among isolates.

### Evaluation of chromogenic medium for rapid identification

Chromogenic culture was evaluated as a rapid and cost-effective method for the presumptive identification of *C. viswanathii*. Isolates previously identified by MALDI-TOF MS were streaked onto CHROMagar™ Candida medium (BD, Franklin Lakes, NJ, USA) alongside reference strains of *C. albicans* and *C. tropicalis*. Plates were incubated aerobically at 30 °C and examined at 24, 48, and 72 h to assess colony morphology and chromogenic characteristics. Colony appearance was documented photographically (Fig. 1) to determine whether *C. viswanathii* exhibited a consistent and distinctive color pattern compared with other *Candida* species.

**Figure 1.**
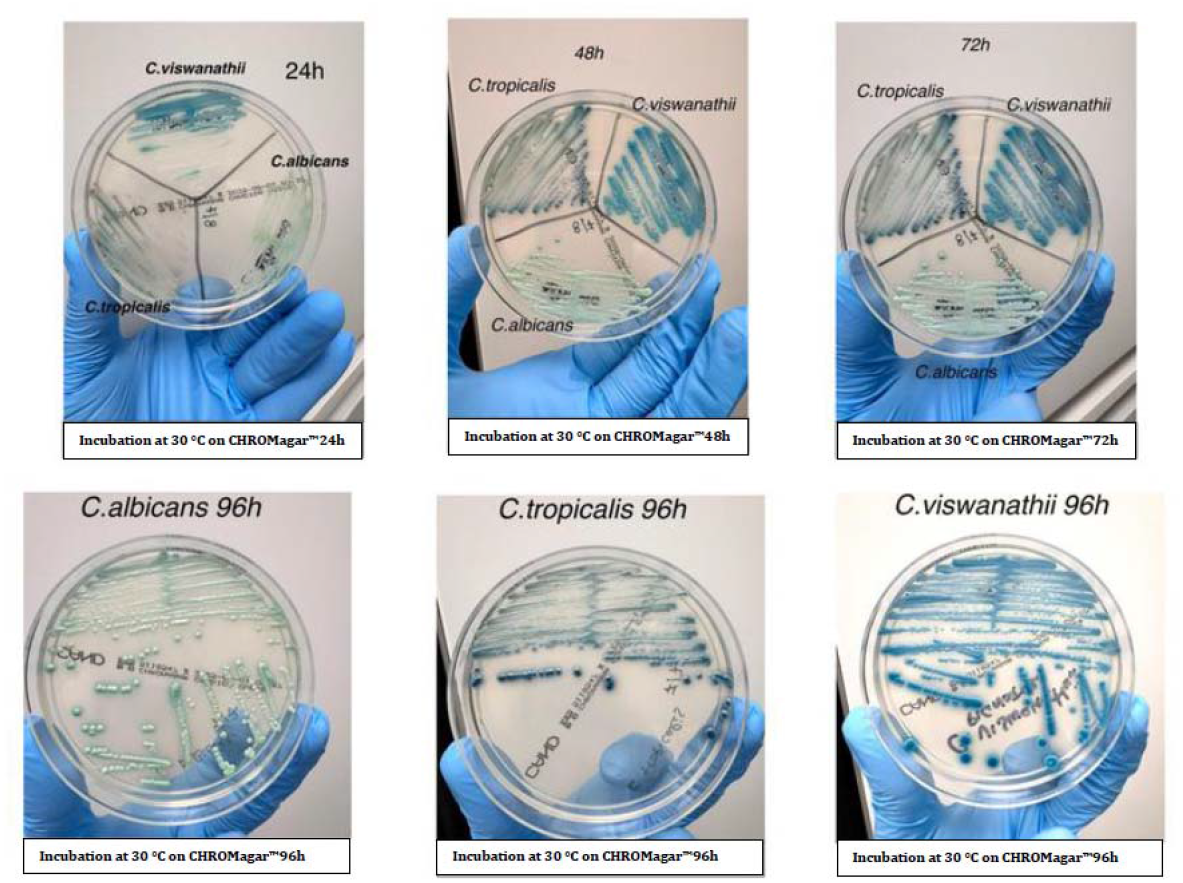
Colony morphology of *Candida viswanathii* on CHROMagar™ Candida medium after incubation at 30 °C for 24 h, 48 h, and 72 h. Colonies developed a characteristic deep blue coloration, distinct from the blue-green hue of C. tropicalis and the light green coloration of C. albicans, supporting the use of this medium as a rapid, low cost preliminary screening tool.

### Genomic DNA extraction

Genomic DNA was extracted from *C. viswanathii* isolates grown for 48 h at 30 °C on Sabouraud dextrose agar plates (Kima, Padua, Italy), as prolonged incubation was used to maximize DNA yield. Colonies were suspended in distilled water, and the fungal pellet was treated with 200 µL of lyticase solution (10 U/mL in G2 buffer supplied with the EZ1&2 DNA Tissue kit) at 37 °C for 90 min with intermittent mixing to promote spheroplast formation. After centrifugation, the pellet was resuspended in 180 µL of the same G2 buffer supplemented with 20 µL of proteinase K and incubated at 56 °C for 15 min. DNA extraction was then performed using the EZ1 automated system (Qiagen BioRobot EZ1, Qiagen, Hilden, Germany) with the EZ1&2 DNA Tissue kit, following the manufacturer’s protocol for bacterial DNA and using an elution volume of 50 µL.

### Whole Genome Sequencing analysis

Genomic libraries were prepared with the Illumina DNA Prep kit and sequenced on a NextSeq 550 platform (2 × 150 bp paired-end). Reads were adapter-trimmed and quality-filtered (Phred >28) using Fastp, with quality control performed by FastQC. Taxonomic assignment and contamination screening were carried out with Kraken2 against the standard fungal database. After trimming, 7.9 million reads were assembled de novo with Shovill, and assembly quality assessed by QUAST. The draft genome comprised 1758 contigs, the largest 187 kb, with a total size of ~25 Mb and GC content of 44.8%. BLAST analysis of the ITS region confirmed 100% identity with Candida viswanathii ATCC 22981.

To confirm species placement, 32 complete Candida genomes from NCBI were analyzed using the UFCG pipeline, and a maximum likelihood phylogeny was reconstructed with IQTREE (1000 bootstraps; best-fit model by ModelFinder). Genomic relatedness with C. tropicalis was evaluated by Average Nucleotide Identity (ANI) using fastANI. Putative azole resistance mutations were screened in ERG1, ERG3, ERG11, MRR1, and UPC2 genes by read mapping to the *C. viswanathii* ATCC 20962 genome (bwa mem) and translation with EMBOSS transeq.

## Results

### Identification of *Candida viswanathii* isolates

A total of fifteen yeast isolates were obtained from positive blood cultures of eight pediatric patients admitted to the NICU (n = 5) and PICU (n = 9) between April and July 2025. Conventional identification by MALDI-TOF MS on both pellets (directly from positive blood cultures) and subcultured colonies produced results consistent with *C. tropicalis* for all isolates. In contrast, the BCID2 panel failed to detect any organism in all cases.

The first isolate collected (patient 1) was subjected to WGS to confirm the species, which unequivocally identified the strain as *C. viswanathii* (Figure S1). This sequenced genome served as the reference for subsequent analyses that confirmed the preliminary results.

### Confirmation of outbreak and strain relatedness by FT-IR spectroscopy

Following the identification of the index isolate, all fifteen isolates were analyzed using FT-IR spectroscopy. Spectra were compared to the reference profile derived from the sequenced isolate. FT-IR analysis showed a high degree of spectral similarity among the eight isolates, forming a tight, distinct cluster with minimal intra-cluster variability. These findings strongly suggest that all isolates belonged to the same outbreak clone (representative dendrograms and LDA plots are shown in Supplementary Figure S2 and S3).

### Chromogenic medium colony appearance

All fifteen isolates displayed a reproducible colony morphology on CHROMagar™ Candida plates. Colonies of *C. viswanathii* developed a distinct deep blue coloration after 24–48 h of incubation at 30 °C, which was maintained at 72 h. This coloration was consistent across all *C. viswanathii* isolates and was visually distinct from the blue-green hue typically produced by *C. tropicalis* and the light green coloration of *C. albicans* under the same conditions (Figure 1). Based on these observations, the chromogenic medium proved useful as a rapid and cost-effective preliminary screening tool for *C. viswanathii*.

### Comparison of MALDI-TOF MS and chromogenic medium performance

Despite the consistent and species-specific color phenotype on Candida ID medium, MALDI-TOF MS was unable to reliably distinguish *C. viswanathii* from *C. tropicalis*. Low discrimination scores (1.6–1.7) were consistently reported, regardless of whether direct identification from the pellet or identification from cultured colonies was performed. These results highlight the current limitation of commercial MALDI-TOF MS libraries for this rare species. A comparison of phenotypic, genotypic, and antifungal susceptibility characteristics of *C. viswanathii* and *C. tropicalis* is presented in Table 1 highlighting the main differences for both laboratory identification and early therapeutic decision-making.

**Table 1.** Key phenotypic, genomic, and antifungal susceptibility differences between *Candida viswanathii* and *Candida tropicalis*.

| Feature | *Candida viswanathii* | *Candida tropicalis* |
| --- | --- | --- |
| Colony color on CHROMagar™ *Candida* | Deep blue after 24–48 h, stable at 72 h | Typical blue green |
| MALDI-TOF MS identification with commercial library | Frequently misidentified as *C. tropicalis* (score 1.6–1.7) | Correctly identified, score $\geq 2.0$ |
| BioFire® BCID panel result | Not detected (no target) | Correctly detected as *C. tropicalis* |
| Genome size (assembly size from WGS) | ~24.2 Mb | ~14.6 Mb |
| Average Nucleotide Identity | 78.12% (38% genome alignable) | — |
| Fluconazole MIC pattern | Frequently elevated, often above epidemiological cutoff values | Generally susceptible |
| Other antifungal agents | Susceptible to echinocandins and amphotericin B | Similar pattern, but usually more susceptible to azoles |
| Clinical frequency | Rare; sporadic cases and limited outbreaks reported | Common; among the top five causes of candidemia |
| Risk of misidentification | High without WGS, FT-IR, or updated MALDI-TOF database | Low |

### Development of an internal MALDI-TOF reference spectrum for *Candida viswanathii*

After WGS confirmed the index isolate as *C. viswanathii* and FT-IR clustered subsequent strains, a local MALDI-TOF MS procedure was implemented to improve identification. Initial analyses of blood culture pellets (Bruker Sepsityper® kit, MBT Smart MALDI-TOF, Compass HT IVD v5.2.330) yielded low discrimination scores (1.6–1.7), often ambiguously matching *C. tropicalis*. To address this, four WGS-confirmed isolates were processed according to the Bruker “Creation of Library Entries” protocol. High-quality spectra were acquired in triplicate, calibrated with BTS, and processed with flexControl/Compass flexAnalysis to generate validated Main Spectrum Projections (MSPs). Incorporation of these MSPs into the local Bruker Taxonomy significantly improved species discrimination, consistently producing scores ≥2.0 for *C. viswanathii* and enhancing routine diagnostic accuracy.

### Genomic relatedness

The WGS data of the index isolate and the spectral clustering results together support that the eight bloodstream isolates represent a single *C. viswanathii* outbreak clone. The close genomic relatedness between *C. tropicalis* and *C. viswanathii* likely explains the misidentification observed with conventional MALDI-TOF MS platforms. Nonetheless comparative genomic analysis revealed a marked difference in genome size between *C. tropicalis* (14.6 Mb) and *C. viswanathii* (24.2 Mb). This divergence was also further supported by the Average Nucleotide Identity (ANI), calculated with fastANI, which yielded an ANI of 78.12% with only 38% of the genome alignable, indicating substantial genomic dissimilarity between the two species (Supplementary Figure S4)

### Genomic characterization of the *Candida viswanathii* outbreak isolates

Phylogenetic and single nucleotide polymorphisms (SNPs) analysis were used to investigate the genetic relatedness among the *Candida viswanathii* isolates from the present outbreak and the two reference genomes available in public databases (NRRL Y-6669 and ATCC 20963). The minimum spanning tree of the first fourteen sequenced isolates showed that all outbreak isolates formed a highly homogeneous cluster, while both reference strains exhibited a high genetic distance (>10,000 SNPs). This findings indicate that the Italian isolates belong to a distinct clonal lineage not represented among currently available *C. viswanathii* genomes.

### Outbreak control measures

Following the confirmation of *C. viswanathii* as the outbreak-causing species, a multidisciplinary infection-control task force was activated. Immediate measures included reinforcement of hand hygiene compliance and environmental cleaning with antifungal-effective disinfectants. Empirical antifungal prophylaxis and treatment regimens were reassessed in collaboration with the Infectious Diseases Unit. Surveillance cultures and environmental sampling were performed to identify potential reservoirs, although no environmental or colonization sources were detected.

The combination of enhanced microbiological vigilance, strict contact precautions, and prompt recognition of the correct pathogen enabled the implementation of an appropriate antifungal regimen, liposomal amphotericin B combined with an echinocandin, which resulted in microbiological eradication and a subsequent slowdown of patient-to-patient (intra-hospital) transmission.

The main challenge encountered was the initial diagnostic delay caused by misidentification through conventional systems; this hindered the timely initiation of effective therapy and allowed early transmission to occur before the pathogen was correctly identified.

## Discussion

This study documents the first outbreak of *C. viswanathii* in Europe, detected at the Bambino Gesù Children’s Hospital in Rome. Fifteen bloodstream isolates from pediatric patients were initially misidentified as *C. tropicalis* by MALDI-TOF MS and were not detected by the FilmArray BCID2 panel, despite microscopic evidence of yeasts. Whole-genome sequencing of the index isolates and subsequent FT-IR spectroscopy confirmed *C. viswanathii* and revealed a clonal cluster responsible for the outbreak. These findings highlight the diagnostic difficulties associated with this rare pathogen and demonstrate the value of an integrated, multimodal approach.

Reports of *C. viswanathii* have historically been restricted to Asia and South America, with the largest case series from India describing 23 bloodstream infections over four years. Our findings extend the geographical distribution of this species to Europe for the first time. Previous studies have emphasized frequent fluconazole resistance and diagnostic challenges due to phenotypic overlap with *C. tropicalis*. Our results confirm both issues and underscore how reliance on conventional diagnostic tools may obscure the true epidemiology of *C. viswanathii*.

The main strength of this investigation lies in its comprehensive diagnostic strategy, which combined phenotypic culture characteristics, MALDI-TOF MS interpretation, chromogenic media, FT-IR spectroscopy, and confirmatory whole-genome sequencing. This multimodal approach enabled early outbreak recognition and species-level confirmation. Limitations include the single-center design, the relatively small number of cases, and the lack of species-specific EUCAST or CLSI breakpoints, which restricted antifungal susceptibility interpretation to epidemiological cut-off values. Furthermore, our retrospective review could not exclude the possibility of undetected cases before 2025 or outside our institution.

Our findings underscore the clinical importance of precise species-level identification in invasive candidiasis. Misidentification of *C. viswanathii* as *C. tropicalis* could have led to inappropriate fluconazole therapy, prolonged fungemia, and a higher risk of dissemination. In contrast, accurate recognition guided the use of amphotericin B or echinocandins in accordance with pediatric guidelines and enabled rapid implementation of infection-control measures, thereby containing nosocomial spread.

The consistently elevated fluconazole MICs observed are consistent with previous descriptions of *C. viswanathii*. Genomic investigations suggest that resistance may be mediated by mutations in ergosterol biosynthesis and regulatory genes such as *ERG11, MRR1*, and *UPC2*. Although our study included preliminary genomic screening for such mutations, further functional validation is required to fully elucidate resistance mechanisms and their potential impact on therapy.

The genomic analysis revealed that the Italian outbreak isolates were clearly distinct from the two publicly available *C. viswanathii* reference genomes (NRRL Y-6669 and ATCC 20963), suggesting the emergence of an independent clonal lineage (Supplementary Fig. S5). The marked genetic distance supports the hypothesis that this outbreak was caused by a previously undescribed genotype, underlining the limited genomic diversity currently represented in public databases and highlighting the need for broader genomic surveillance of *C. viswanathii*.

Although extensive microbiological and epidemiological investigations were conducted, the route of *C. viswanathii* introduction into Italy remains unresolved. To date, all environmental samples and rectal swabs collected from affected patients have tested negative, leaving the source of the outbreak elusive. Potential routes still under consideration include contamination of medical devices, pharmaceuticals administered during treatment, or hospital water systems and inanimate surfaces acting as transient reservoirs for this rare yeast.

Future studies should assess the prevalence of *C. viswanathii* across Europe and beyond, as its apparent rarity may reflect underrecognition. Multicentre surveillance initiatives are needed to clarify its epidemiology, while continuous updates of MALDI-TOF MS reference libraries and syndromic panels will be essential to ensure accurate species identification. In parallel, genomic investigations are required to elucidate antifungal resistance mechanisms and the evolutionary relationship between *C. viswanathii* and *C. tropicalis*. Together, these efforts will enhance diagnostic accuracy, guide antifungal stewardship, and reinforce infection-prevention strategies for emerging fungal pathogens.

From a European public health perspective, this outbreak represents the first documented emergence of *C. viswanathii* in Europe and underscores the need for continent-wide genomic surveillance of rare fungal pathogens. Key lessons include the value of rapid molecular confirmation in atypical *Candida* clusters, the integration of FT-IR spectroscopy for early outbreak recognition, and the use of whole-genome sequencing to confirm species identity and strengthen diagnostic preparedness through continuously updated databases. Moreover, this experience highlights how close collaboration among clinical, microbiology, and infection-control teams can ensure timely containment of fungal outbreaks, even in the absence of a clearly identified environmental source.

## Data Availability

All data produced in the present work are contained in the manuscript

## Author Contributions

G.V., V.C., V.F., C.F.P., and P.B. contributed to the conceptualization of the study. G.V., V.C., V.F., G.F., S.R., M.C., and S.C. contributed to the methodology. G.V., V.C., V.F., C.F.P., and P.B. performed the formal analysis. G.V., V.C., V.F., G.F., S.R., M.C., B.L., M.O., S.C., M.R., M.P.R., A.D., A.C., A.V., L.G., L.D.C., and M.L.C.D.A. contributed to the investigation. G.V., V.C., V.F., C.F.P., and P.B. prepared the original draft. G.V., V.C., V.F., M.R., M.Ra., G.F., C.F.P., and P.B. contributed to the review and editing of the manuscript. C.F.P. and P.B. supervised the study. C.F.P. was responsible for funding acquisition.

All authors have read and approved the final version of the manuscript.

## Institutional Review Board Statement

“Not applicable” for studies not involving humans or animals.

## Informed Consent Statement

Considering the nature of the analysis, the current study did not require the approval of the local ethics committee according to current legislation and with the 1964 Helsinki Declaration. In addition, parents or legal guardians of patients provided consent to use personal data for diagnosis, treatment and related research purposes at the time of hospitalization.

No human subjects or identifiable human data were involved in our research. The work was performed exclusively on bacterial isolates obtained from residual material of routine blood cultures collected for standard clinical diagnostics in hospitalized pediatric patients. After clinical reporting, the leftover material is routinely discarded, and the isolates are completely anonymized and de-identified, with no link to patient information. For this reason, ethical approval was not required for this study.

## Data Availability Statement

All data generated or analyzed during this study are included in this published article. Further inquiries can be directed to the corresponding author.

## Acknowledgments

The authors thank Tommaso Vito, Maria Del Carmen Pereyra Bozza, and Teresa D’Urbano for their valuable technical assistance and support during the study. The authors also wish to thank all staff of the Microbiology and Virology Laboratory of Bambino Gesù Children’s Hospital IRCCS for their outstanding technical support in processing samples, performing laboratory analyses, and managing data.

