## Supplemental Table 1 for "When Conventional Methods Fail: First Detection of a *Candida viswanathii* Outbreak in Europe in a Paediatric Hospital Revealed by Whole Genome Sequencing and FT-IR Spectroscopy": MIC_Candida_viswanathii_table_S1.pdf

Table S1. Minimum inhibitory concentrations (MICs, mg/L) of antifungal agents against Candida viswanathii isolates from different patients.

| Paziente | Amphotericin B | Anidulafungin | Caspofungin | Fluconazole | Isavuconazole | Itraconazole | Micafungin | Posaconazole | Voriconazole |
|---|---|---|---|---|---|---|---|---|---|
| 1 | 0.5 | 0.06 | 0.06 | 4.0 | 0.25 | 0.12 | 0.06 | 0.25 | 0.25 |
| 2 | 1.0 | 0.25 | 0.12 | 4.0 | 0.5 | 0.25 | 0.12 | 0.25 | 0.5 |
| 3 | 0.5 | 0.12 | 0.12 | 4.0 | 0.5 | 0.25 | 0.06 | 0.25 | 0.5 |
| 4 | 0.5 | 0.12 | 0.12 | 4.0 | 0.5 | 0.25 | 0.06 | 0.25 | 0.5 |
| 5 | 0.5 | 0.12 | 0.06 | 4.0 | 0.5 | 0.12 | 0.06 | 0.25 | 0.5 |
| 6 | 0.5 | 0.12 | 0.25 | 4.0 | 0.5 | 0.25 | 0.06 | 0.25 | 0.5 |
| 7 | 0.25 | 0.12 | 0.12 | 4.0 | 0.5 | 0.12 | 0.03 | 0.12 | 0.5 |
| 8 | 0.25 | 0.12 | 0.12 | 4.0 | 0.5 | 0.12 | 0.06 | 0.25 | 0.5 |
| 9 | 0.25 | 0.12 | 0.12 | 4.0 | 0.25 | 0.12 | 0.03 | 0.25 | 0.25 |
| 10 | 0.25 | 0.12 | 0.12 | 2.0 | 0.5 | 0.12 | 0.06 | 0.25 | 0.25 |
| 11 | 0.5 | 0.12 | 0.12 | 2.0 | 0.5 | 0.12 | 0.06 | 0.25 | 0.25 |
| 12 | 0.25 | 0.12 | 0.12 | 2.0 | 0.5 | 0.12 | 0.06 | 0.25 | 0.5 |
| 13 | 0.25 | 0.12 | 0.12 | 4.0 | 0.5 | 0.12 | 0.06 | 0.25 | 0.5 |
| 14 | 0.5 | 0.12 | 0.12 | 2.0 | 0.5 | 0.12 | 0.06 | 0.25 | 0.25 |
| 15 | 0.25 | 0.12 | 0.12 | 2.0 | 0.5 | 0.12 | 0.06 | 0.25 | 0.25 |
