## Supplemental Figure 2 for "When Conventional Methods Fail: First Detection of a *Candida viswanathii* Outbreak in Europe in a Paediatric Hospital Revealed by Whole Genome Sequencing and FT-IR Spectroscopy": Figura S2.pdf

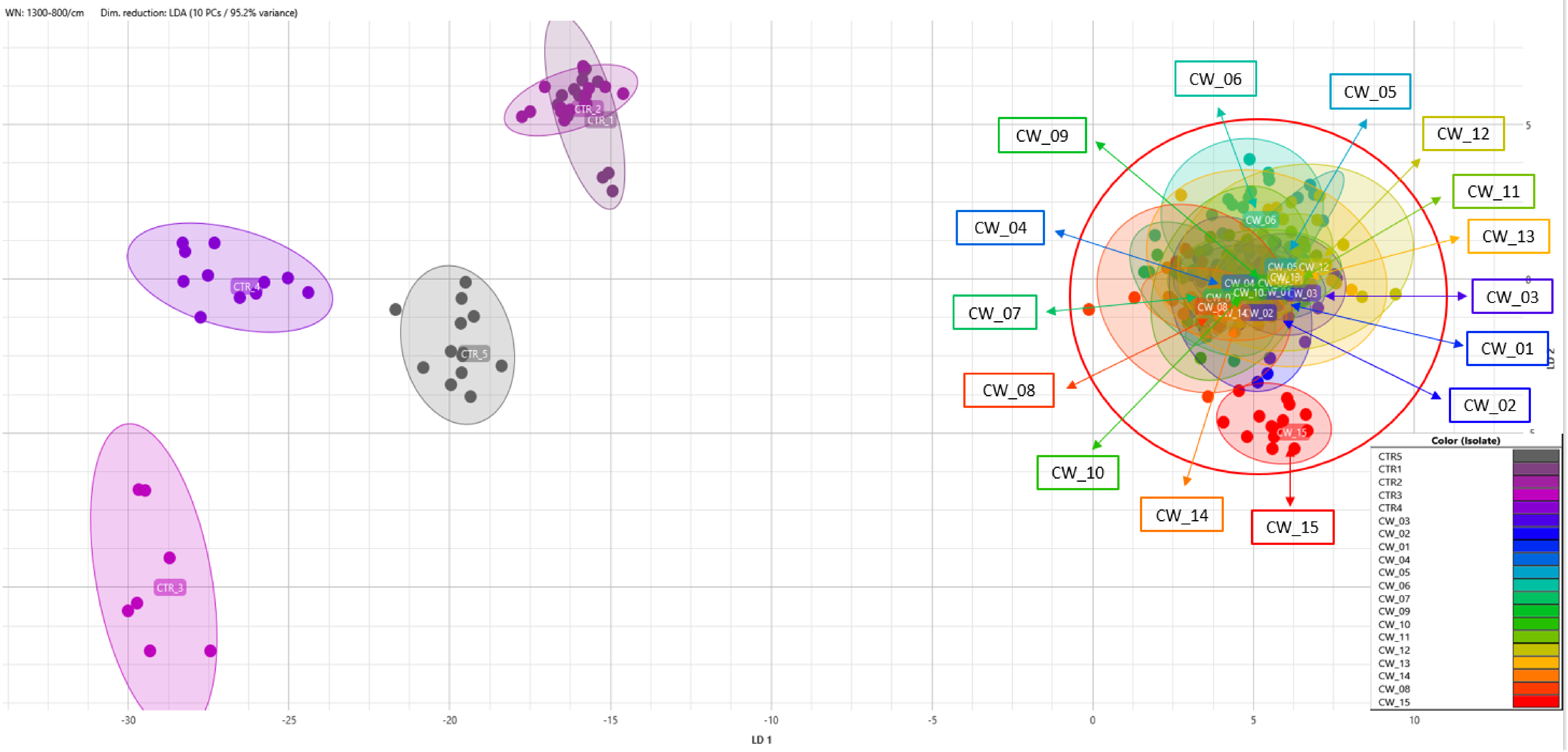


**Figure S2.** Scatter plot generated by FT-IR Bruker analysis (spectral range 1300–800 cm$^{-1}$; dimensionality reduction by LDA using 9 principal components, explaining 95.7% of the variance) showing the *Candida viswanathii* outbreak isolates clustering together. Four external *Candida* strains, used as controls, are clearly separated into distinct groups. Ellipses represent 95% confidence intervals for each cluster.
