## Supplemental Figure 3 for "When Conventional Methods Fail: First Detection of a *Candida viswanathii* Outbreak in Europe in a Paediatric Hospital Revealed by Whole Genome Sequencing and FT-IR Spectroscopy": Figura S3 (1).pdf

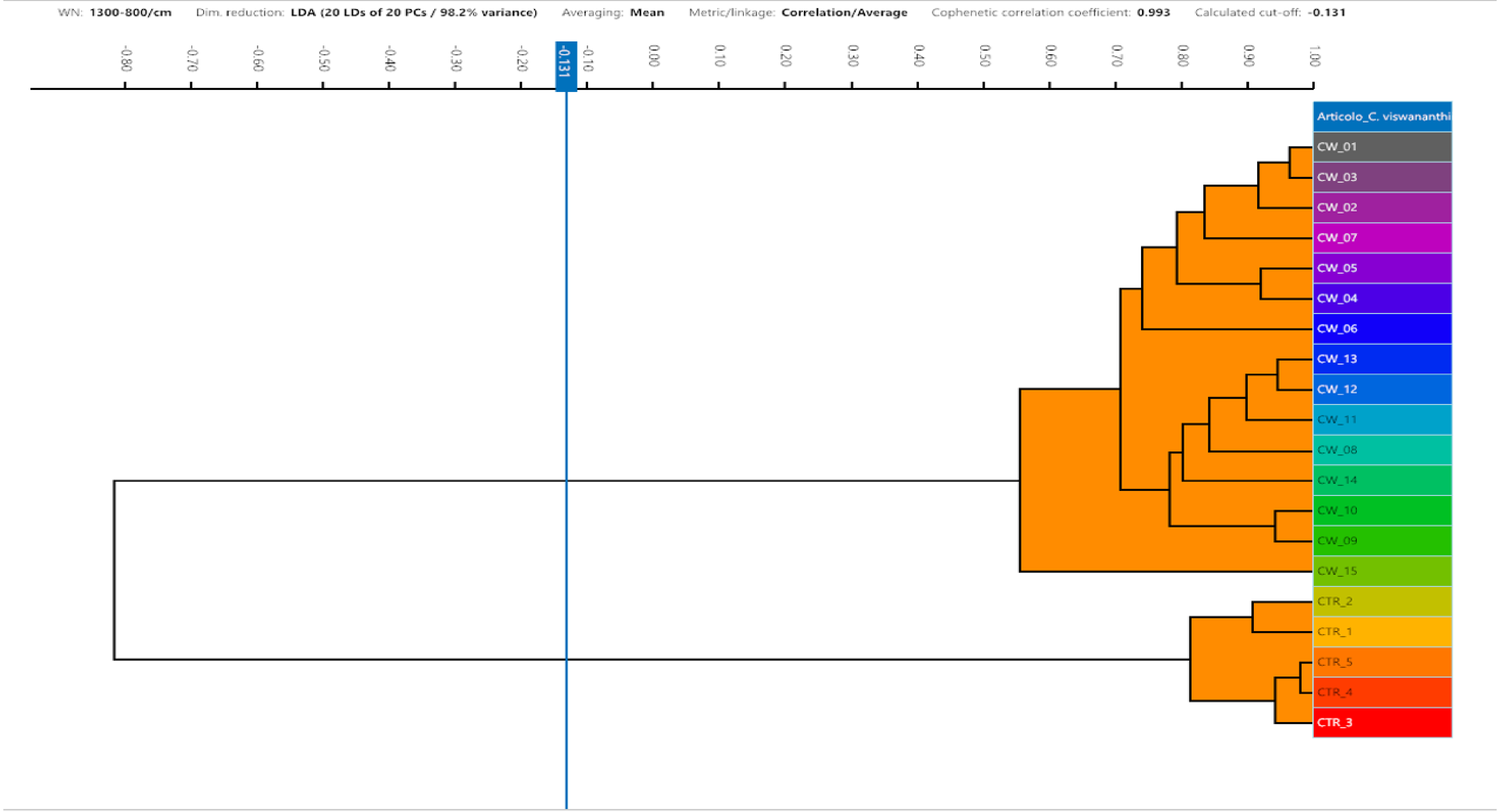


**Figure S3.** Dendrogram generated by FT-IR Bruker analysis (spectral range 1300–800 $cm^{-1}$; dimensionality reduction by LDA using 18 linear discriminants from 20 principal components, explaining 98.7% of the variance) showing the clustering of *Candida viswanathii* outbreak isolates together. Four external *Candida* strains, used as controls, are clearly separated from the outbreak cluster. The analysis was performed using the Euclidean distance metric and average linkage, with a cophenetic correlation coefficient of 0.933 and a calculated cut-off value of 29.51.
