## Supplemental Figure 4 for "When Conventional Methods Fail: First Detection of a *Candida viswanathii* Outbreak in Europe in a Paediatric Hospital Revealed by Whole Genome Sequencing and FT-IR Spectroscopy": Figure S4.pdf

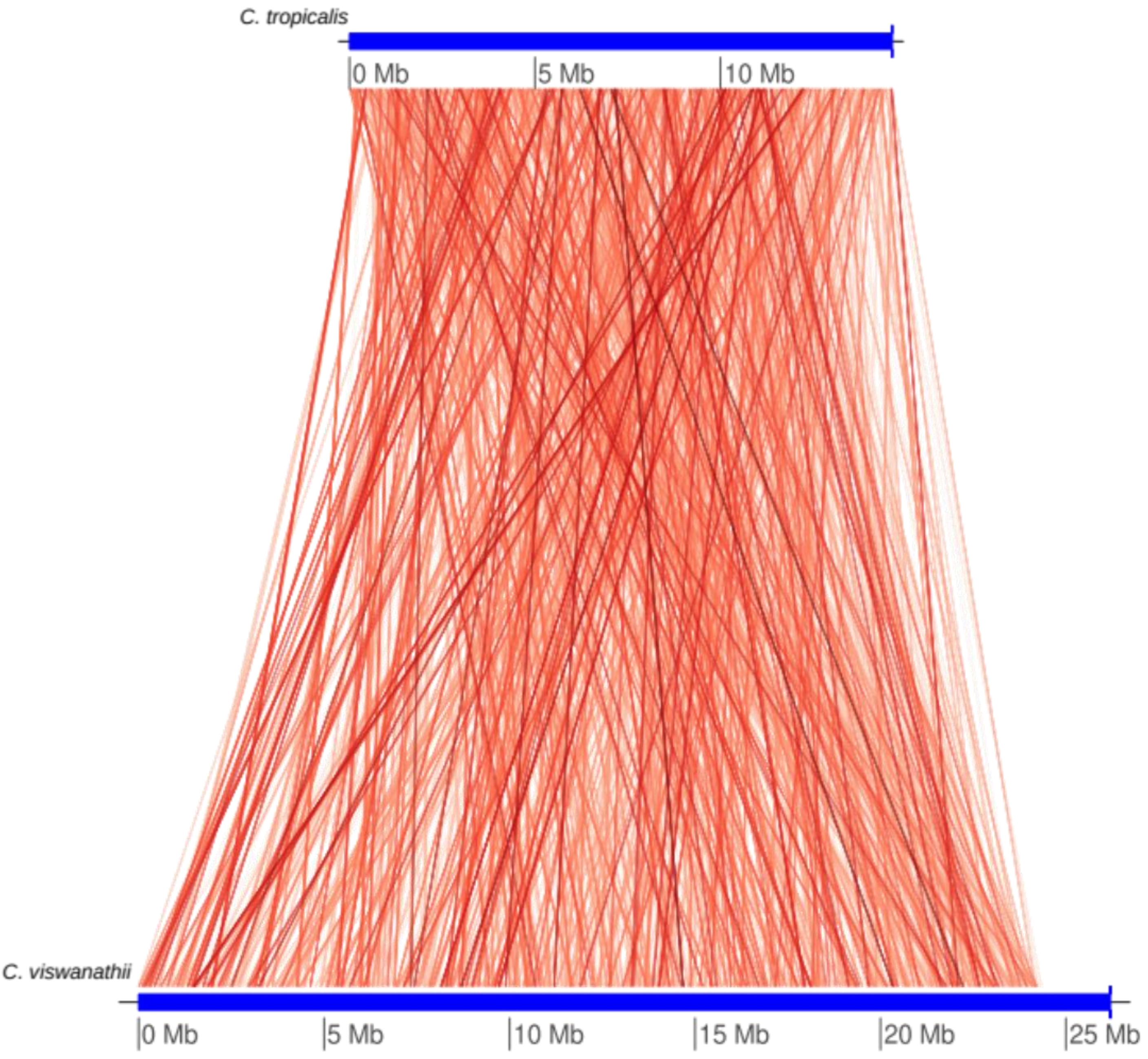


**Figure S4. Visualization of conserved region between *C. tropicalis* and *C. viswanathii* (sample CW_01) genomes by fastANI**
