## Supplemental Figure 5 for "When Conventional Methods Fail: First Detection of a *Candida viswanathii* Outbreak in Europe in a Paediatric Hospital Revealed by Whole Genome Sequencing and FT-IR Spectroscopy"

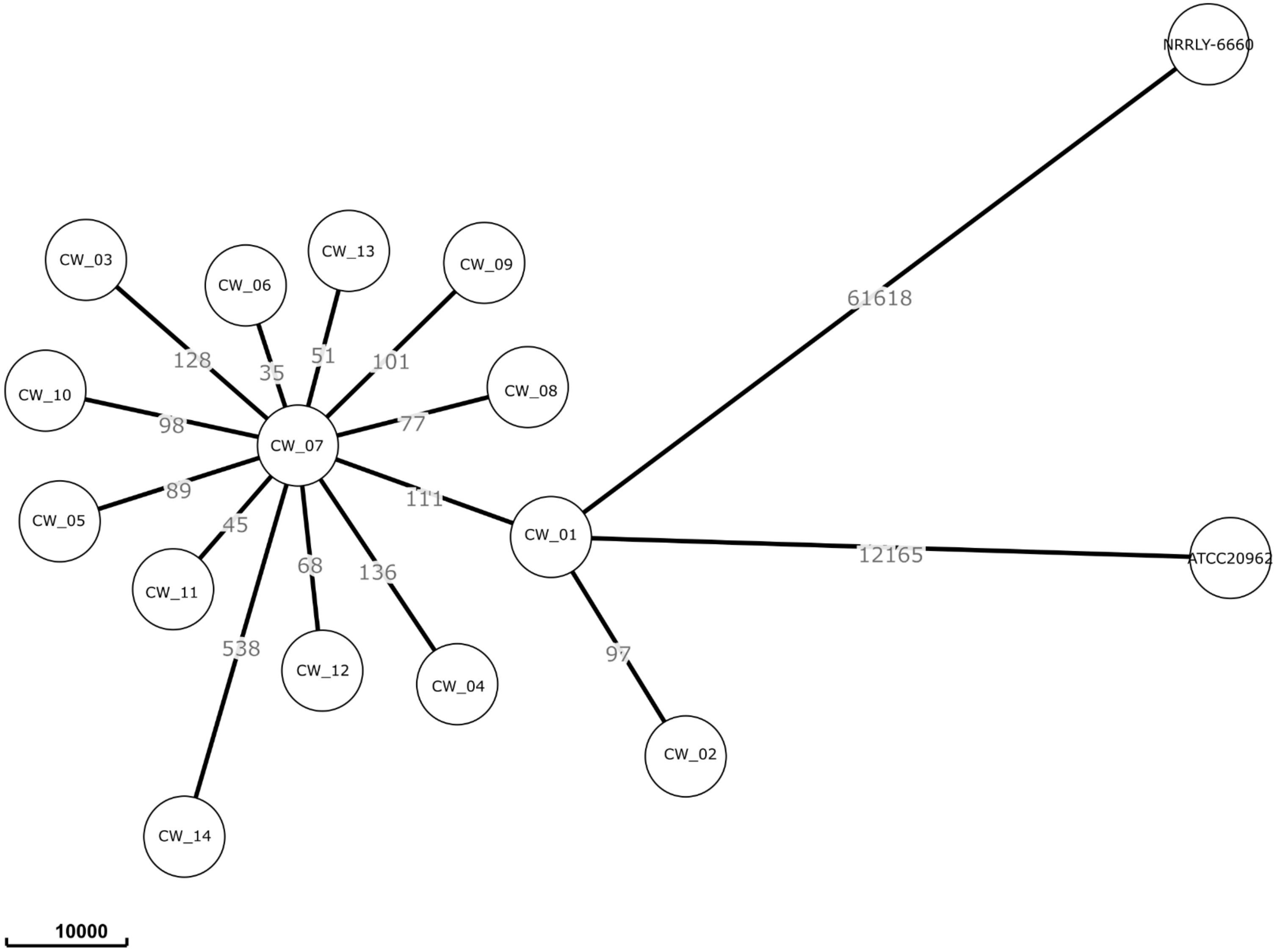


**Figure 5.** *Minimum spanning tree (WGS-SNP analysis) of* **Candida viswanathii** *outbreak isolates.*
All Italian isolates formed a highly homogeneous cluster, consistent with a single clonal lineage. Reference strains NRRL Y-6669 and ATCC 20963 were separated by >10,000 SNPs, indicating marked genomic divergence and supporting the emergence of a distinct lineage.
